# Conditional Optimism and Systemic Constraints: Healthcare Provider Perceptions of AI in Safety-Net Settings Using a Mixed-Methods Vignette Approach

**DOI:** 10.64898/2026.09.29.26364281

**Authors:** Ishani Purohit, Matt Kammer-Kerwick, Emily Spandikow, Gregory Pogue, S. Craig Watkins

## Abstract

As health AI tools enter clinical spaces, how they are perceived by providers in safety-net environments shapes whether their benefits reach underserved populations. We examined the degree and drivers of healthcare providers’ trust in health AI, whether that trust moderates perceived benefits for safety-net populations, and how providers balance benefit against concern across four specific hypothetical AI tools. We used a two-study mixed-methods design: 18 semi-structured interviews with healthcare practitioners and research leaders, followed by a survey of 229 Texas healthcare providers incorporating four vignettes describing hypothetical AI tools, each rated for appeal and concern and followed by open-ended explanation. Responses were thematically coded and stratified by self-reported trust in health AI (47.6% high trust, 52.4% low trust). Low-trust respondents most often cited building trust (50.5%) and data bias and accuracy (33.0%) as drivers of their current trust level, while high-trust respondents more often cited efficient and effective care (23.5% vs. 3.7% low-trust, p<0.001) and accuracy of data (11.8% vs. 1.8% low-trust, p=0.004); concerns about data security, governance, and the need for human oversight were raised at similar rates by both groups. When asked how AI could specifically benefit safety-net populations, both groups converged on expanding access to care (45.9% each), and none of 25 benefit-related codes differed significantly by trust group. Across all four vignettes, mean appeal ratings exceeded mean concern ratings, and high-trust providers rated every tool as more appealing and, for three of four tools, less concerning, without changing which tool ranked where relative to others. Providers exhibit conditional optimism: support is contingent on governance, human oversight, and context-sensitive implementation rather than on the technology itself. Realizing AI’s promise for safety-net populations will require human-in-the-loop models, localized implementation strategies, community-informed design, and engaging providers directly in policy design and tool development.

**Author Summary:** Artificial intelligence is increasingly built into the everyday tools that healthcare workers use, but whether it actually helps the patients who need the most support depends on what those workers think of it and how much they trust it. We talked with 18 healthcare workers and researchers, then surveyed 229 healthcare providers across Texas, many of whom work in clinics and hospitals that serve low-income and rural communities. About half of the providers we surveyed trusted health AI and half did not. Providers who trusted AI less pointed to a need to build trust and worries about biased or inaccurate data; providers who trusted it more pointed to gains in efficient, accurate care. Both groups agreed, in similar numbers, that AI’s biggest promise for underserved patients is expanding access to care, and neither group described that promise differently from the other. When we showed providers four realistic AI tools, all of them found more to like than to worry about. Our findings suggest that providers are not simply for or against health AI. Their support depends on whether AI is deployed with real human oversight, sound governance, and attention to the specific setting where it is used.

## Introduction

The rapid integration of artificial intelligence into healthcare delivery raises critical questions about equity, trust, and feasibility in safety-net settings. While AI is often framed as a mechanism for improving access and efficiency, the perspectives of providers serving underserved populations remain underexamined. Safety-net populations are defined as individuals and communities characterized by limited access to healthcare services, including those who are uninsured or underinsured, socioeconomically disadvantaged, or served by resource-constrained systems. We employ a mixed methods approach to study the perceptions of health care providers relative to the promises of responsible deployment of AI and the potential perils that need to be navigated to achieve that goal. Specifically, we employed a two-study design that involved a set of exploratory interviews among thought leaders and practitioners followed by a survey of practitioners to broaden the perspectives included in our analysis. We chose to focus on safety-net populations because of the unique challenges faced by healthcare providers in serving those populations, including high patient-to-provider ratios, significant constraints on resources, and greater health disparities than in the general population. This study is part of a broader research initiative on health AI adoption in safety-net and rural health care settings, portions of which were first described in a 2025 institutional report (1).

The survey used in this exploratory study included both quantitative and qualitative questioning; our focus in this paper is on the qualitative insights obtained. It is important to note that the survey design included a series of structured vignettes that presented health care providers with specific realistic but fictional health AI technologies. Within these vignettes, healthcare practitioners provided ratings for the degree of appeal and the degree of concern associated with deploying the described technology among safety net community members. The survey also asked healthcare providers to provide a qualitative explanation of how they were thinking about responsible deployment of that technology in the context of those perceived points of appeal and points of concern.

We performed a literature review that allowed us to focus on specific gaps and used the interviews to develop a more structured line of questioning for the survey. From this first phase we established the following research questions:

- RQ1: To what degree do healthcare providers trust health AI, and what factors shape that trust?
- RQ2: How might health AI benefit safety-net populations, and does providers’ baseline trust in AI moderate those perceived benefits?
- RQ3: How do healthcare providers balance the benefits of health AI against their concerns, as explored through provider reactions to four specific hypothetical AI tools?

We believe that responses to these questions from the perspectives of safety-net healthcare providers will provide unique insights regarding the design and deployment of health AI in general and in under-resourced settings, specifically.

## Literature Review

Artificial intelligence has the potential to transform healthcare delivery, particularly in rural and underserved settings where resource limitations and workforce shortages create significant barriers to care. These settings, often characterized by high patient-to-provider ratios, fragmented healthcare infrastructure, and limited access to specialty services, stand to benefit greatly from AI-driven solutions aimed at improving diagnostic accuracy, clinical workflows, and patient management (2,3). However, the integration of AI into these environments presents unique technical, financial, and ethical challenges that must be addressed to ensure equitable access to high-quality care.

Rural safety-net hospitals serve populations that are disproportionately low-income, uninsured, and burdened with chronic diseases, which can complicate care delivery (4). AI-enabled technologies such as predictive analytics, natural language processing, and decision-support systems offer opportunities to streamline clinical workflows, optimize resource allocation, and mitigate disparities in healthcare access (5–7). Studies have shown that AI-based clinical decision support tools can enhance diagnostic accuracy and reduce cognitive overload for clinicians, which is particularly beneficial in low-resource settings where providers often manage a wide range of complex cases with limited specialist support (8). Additionally, the use of AI-driven administrative automation, such as auto-populating clinical notes, processing billing, and managing electronic health records, can significantly reduce the time providers spend on non-clinical tasks (9). This is especially valuable in safety-net settings, where limited staffing and high caseloads often lead to documentation burdens that contribute to provider stress and burnout.

A study incorporating specific AI models for Hispanic and Black women diagnosed with breast cancer shows how specific models can further improve diagnostic and treatment accuracy in underserved and underrepresented populations. These models outperformed general models by better predicting survival outcomes based on their unique health profiles. By tailoring AI models to the unique healthcare needs and disparities of specific demographic groups, these technologies can provide more accurate, individualized care, ultimately improving outcomes and promoting equity (10).

For example, tests with Woebot and Crisis Textline chatbots showed a significant reduction in anxiety and symptom mitigation compared to historical values for human intervention (11). If these benefits can be confirmed, these tools could improve equal access, expand availability in terms of hours per day and user capacity, while offering anonymity and reduced stigma.

Populations whose health suffers from the challenges of access and cost while simultaneously being underserved, uninsured, or have difficulty accessing care due to their location are safety net populations. These populations, including rural communities, have reduced access to quality health care and usually rely on publicly funded services because of economic hardship and systemic inequities while experiencing limited access to health care and social support. Although the specific factors may differ, structural barriers hinder their access to essential services and opportunities for improved health outcomes.

Despite these advantages, health IT adoption in rural and underserved communities has historically lagged due to systemic barriers such as interoperability issues, digital infrastructure limitations, and financial constraints (3,12). Research consistently finds that one of the major barriers to advanced health-based technological solutions in rural areas is inadequate internet connectivity (13–17) And while lower-income households in urban areas have access to the internet primarily via mobile phones, they often encounter struggles to afford consistent access to broadband services (18,19). In both instances, the deployment of AI-based health solutions associated with remote patient monitoring or digital biomarkers face notable challenges.

The implementation of AI in these settings must be approached with a focus on equity, feasibility, and long-term sustainability to avoid exacerbating existing healthcare disparities. AI models trained on predominantly urban and insured populations may fail to generalize to rural patient populations, leading to bias in diagnostic recommendations and clinical predictions (2). Addressing this issue requires deliberate efforts to ensure inclusive and representative training datasets that reflect the sociodemographic and epidemiological profiles of rural and underserved communities (20). A study in Tennessee incorporated social determinants of health (SDOH), such as access to healthcare, transportation, and socioeconomic status, to demonstrate the importance of tailoring AI models to local conditions. Machine learning models that integrated county-level SDOH data showed access to healthcare and vaccination rates were critical in predicting COVID-19 risk and could be applied similarly in rural settings to improve AI predictions and health outcomes. Incorporating community-specific data ensures that AI models are more accurate and reflective of the challenges faced by underserved populations, ultimately promoting equity in care delivery (21).

Additionally, safety-net hospitals and rural clinics often lack the financial resources to invest in AI-driven infrastructure. Unlike for-profit institutions, these hospitals rely heavily on Medicaid and fixed tax revenues, which limits their ability to adopt new technologies without external funding or policy incentives (22). AI solutions for these settings must be cost-effective and easily integrated into existing electronic health records (EHRs) to maximize their impact. Browning et al. (2020) also emphasizes the need for vendor-neutral AI platforms that minimize interoperability challenges, as many safety-net hospitals operate with heterogeneous IT systems due to cost-driven procurement decisions.

Furthermore, patient engagement and digital literacy remain critical factors influencing the success of AI-driven health interventions in rural areas. Studies indicate that racial and ethnic minority populations, who often comprise a significant portion of safety-net hospital patients, use digital health tools at lower rates due to barriers such as lack of trust in AI, poor access to broadband, and limited familiarity with health IT systems (8). A lack of digital literacy among patients and healthcare providers can limit the effective utilization of AI-driven tools (23). AI implementation strategies must therefore include culturally tailored patient education and community engagement efforts to ensure that underserved populations can meaningfully interact with these technologies.

While prior research has examined barriers to AI adoption broadly, there is limited empirical work capturing how frontline providers in safety-net contexts evaluate both the benefits and risks of AI and how they negotiate these trade-offs in practice. A recent systematic review of health-worker trust in clinical AI confirms this heterogeneity more broadly (24), though safety-net-specific contexts remain rarely examined directly.

## Methods

### Study Design Overview

This study utilized a mixed methods approach to explore healthcare professionals’ perceptions of Health AI (HAI) technologies, particularly in the context of safety-net care. The research design included an extensive review of the literature, a health care provider survey, and a series of in-depth interviews with healthcare practitioners and researchers. These components allowed for the triangulation of insights across conceptual, experiential, and practice-based domains.

Both phases of this project were granted an exempt determination by our institution’s IRB (STUDY00005660). Following standard procedures, the survey obtained consent in the first screen of the survey. The interviews verbal obtained consent at the beginning of the survey. Participants who did not consent, respectively, exited the survey or were not interviewed.

The survey component focused on understanding current awareness and general attitudes toward Health AI, as well as reactions to four hypothetical AI tool deployment scenarios. Open-ended responses captured nuanced perspectives about AI’s utility, feasibility, and perceived risks across different clinical and patient contexts. The interviews were used to deepen understanding of themes identified in the literature and survey by eliciting rich, narrative accounts of participants’ real-world experiences and professional insights. All open-ended responses, including those tied to vignettes, were analyzed qualitatively using a shared coding framework.

Study 1 informed the development of Study 2 instruments, including vignette design and survey constructs.

### Study 1: Interviews

To explore provider experiences and perspectives on health AI more deeply, the research team conducted 18 in-depth, semi-structured interviews. Fourteen interviews were conducted with healthcare practitioners, and four with research leaders. The thematic narrative presented below draws primarily on the subset of interviews with organizational and policy leaders; frontline practitioner perspectives are represented throughout Study 2’s survey results. Participants were recruited through the research team’s professional network, including through referrals from health system partners. The interviews emphasized themes related to healthcare delivery in safety-net settings.

Interviews were conducted virtually via Zoom and lasted approximately 30 to 60 minutes. The interview guide included open-ended prompts that addressed a range of topics, such as:

- Levels of trust in AI and factors that enhance or erode that trust
- Perceived benefits and concerns surrounding AI integration in clinical workflows
- Ethical considerations and bias in AI models
- Infrastructure and interoperability challenges
- Patient-provider communication and engagement, especially in low-resource settings
- The role of AI in advancing or undermining health equity

The 2nd and 5th authors developed a sampling frame of 63 potential participants. Interviews were conducted from July 18, 2024, to September 3, 2024, with 14 interviews conducted in July and August. Fourteen participants were based in Texas and 13 had direct experience serving rural and safety-net communities. The 1st, 3rd, 4th, and 5th authors, assisted by a graduate student, conducted the interviews. Potential participants were approached by email; of the 63 individuals in the sampling frame, 45 did not respond to the invitation and were not interviewed. All interviews were transcribed verbatim and anonymized for analysis. Interviews were audio- and video-recorded via Zoom.

Coding was conducted by multiple members of the research team using a shared codebook developed iteratively from initial transcripts and literature-informed constructs, managed using Dedoose. We performed thematic analysis (25) with multiple coders. The 2nd author participated in the thematic analysis with the lead and 4th authors. Coders independently coded subsets of transcripts and met regularly to resolve discrepancies and refine code definitions through qualitative reconciliation. A qualitative coding framework consisting of 66 individual codes was developed to capture sentiments expressed across the interviews. These codes were grouped into broader thematic domains representing both the “promise” and “peril” of AI in healthcare, with minor sub-themes identified within each of the five major thematic domains that emerged. Selective coding was used to highlight particularly illustrative narratives related to safety-net implementation. Quotes from the interviews were later compared and aligned with themes that emerged from the survey data. Illustrative quotations ultimately selected for presentation in Results were checked against the full coded dataset to confirm their representativeness of the broader thematic patterns. The entire team iteratively reviewed the interview data and reflected on the codes and themes that emerged. Interviewing was extended until the team judged that thematic saturation had been reached.

A completed COREQ (Consolidated Criteria for Reporting Qualitative Research) checklist for the Study 1 interviews is provided in the Supplement.

### Study 2: Surveys

The second phase of data collection involved a statewide provider-facing survey designed to capture healthcare professionals’ perceptions of health AI, particularly in the context of rural and underserved populations. The survey was piloted in August 2024 within the research team’s healthcare provider network and subsequently fielded through Dynata, a healthcare panel provider, from September 5 to September 24, 2024. A total of 229 complete responses were collected from healthcare providers across Texas, representing a variety of roles and healthcare settings. Eligibility was determined by two upfront screening questions confirming that the respondent’s organization operated in an eligible Texas Health and Human Services region and served at least one specified safety-net-relevant patient population (e.g., low-income, uninsured, rural, or otherwise underserved groups); respondents who did not meet either criterion were not permitted to continue.

A completed CHERRIES (Checklist for Reporting Results of Internet E-Surveys) checklist for the Study 2 survey is provided in the Supplement.

Study 2 included semi-structured lines of questioning about how participants’ thought about their current level of trust in AI technologies to support healthcare delivery

While the majority of the survey consisted of quantitative items, several open-ended questions invited participants to elaborate on key topics. These questions were positioned directly after scaled questions and functioned as semi-structured qualitative data, offering context and rationale behind respondents’ ratings. The general open-ended questions included:

- “In a sentence or two, please describe what impacts your level of trust in AI technologies to support healthcare delivery.” *(Linked to question which asked respondents to rate their current level of trust in HAI)*
- “Please list two ways you think AI could specifically benefit rural and other safety-net populations in healthcare.”

We performed thematic analysis (25) with multiple coders and have focused our analysis of emergent themes as appropriate by level of trust. Our analysis of these topics includes an overall view of the sample plus a stratification by current level of trust (low vs high). Specifically, respondents’ level of trust was measured using a 5-point Likert scale response to “To what extent do you trust AI technologies to support healthcare delivery?” where 1 = completely trust, 2 = somewhat trust, 3 = Neutral, 4 = somewhat distrust, and 5 = completely distrust. (note that this item is reverse-coded relative to the appeal and concern scales below). Participants were classified as high trust if they selected the upper end of the scale (1 or 2, i.e., “completely trust” or “somewhat trust”) and low trust if they selected the lower end of the scale (3, 4, or 5, i.e., “neutral,” “somewhat distrust,” or “completely distrust”). Neutral responses were grouped with distrust rather than trust because the analytic contrast of interest is between providers who affirmatively extend trust to health AI and those who withhold it, whether through skepticism or through undecidedness. We note that this makes the low-trust group heterogeneous: 73 of its 120 members (60.8%) selected “neutral” rather than a distrusting response.

In addition to these questions, the survey also included four health AI vignettes, using an approach similar to (26). The survey assessed the appeal and concern associated with specific Health AI applications. Each vignette described a hypothetical AI tool for clinical use among safety-net populations. The technologies chosen for the vignettes were designed based on needs and interests expressed in the literature.

The vignettes presented the opportunity to move from abstract discussion to more specific applications of AI in healthcare. See Methods and S1 Text for the full vignette descriptions.

The vignettes were developed based on themes identified in the literature review and findings from Study 1 interviews, ensuring that each scenario reflected realistic AI applications relevant to safety-net care contexts.

`o` DocuScribeAI is an AI tool designed to assist in transcribing and organizing clinical notes during patient interactions.
`o` TherapiaAI analyzes real-time data from patient histories, genetic information, and current health conditions. It suggests customized therapeutic plans based on clinical guidelines and patient-specific factors.
`o` HealthRiskAI integrates patient-specific social determinants of health and provides real-time risk assessments.
`o` CommCare analyzes communication preferences and engagement patterns, offering insights to improve patient-provider interactions.

DocuScribe draws from AI-powered transcription technologies, particularly large language models (LLMs), to automate clinical documentation and ease administrative burden. These tools generate structured notes from physician–patient interactions, improving workflow efficiency and reducing burnout, while enabling more patient-centered care (27). However, risks such as omissions, fabrications, misattributions, and limited contextual understanding, especially around nonverbal cues and speaker differentiation, can compromise accuracy (28).

TherapiaAI is inspired by AI tools in personalized medicine that use genomic, clinical, and lifestyle data to generate individualized treatment plans. These applications enhance diagnostic precision and enable earlier interventions, especially in complex or ambiguous cases, by synthesizing diverse health inputs. Yet, they also raise concerns around biased data, privacy of sensitive genomic information, and clinician challenges in interpreting complex AI outputs due to limited training and infrastructure (29,30).

HealthRiskAI builds on AI systems that incorporate patient-specific social determinants of health (SDOH) to improve diagnosis and care navigation (21). These models dynamically integrate factors such as poverty, transportation access, and vaccination rates to produce real-time risk forecasts and guide equitable care coordination (31). For these systems to fulfill their promise, designers must ensure bias mitigation, interoperability of data, and transparency in how AI-generated risk insights are used in clinical workflows (32).

CommCare was inspired by AI tools that analyze communication preferences and engagement behaviors to improve patient-provider interactions. Using data from EHRs, portals, and care platforms, these tools identify how patients prefer to receive information (e.g., SMS vs. calls) and can flag early signs of disengagement, prompting proactive outreach (33). By aligning communication strategies with patient-specific behaviors, these systems enhance adherence, satisfaction, and trust in care relationships (34). While, AI-drafted patient portal replies to patient queries reduced clinician workload, errors have occurred, often submitted unedited (35).

For each vignette, respondents were asked to rate the tool’s appeal and potential concern on a Likert scale, followed by an open-ended prompt: “In a sentence or two, tell us a little more about the appeal you perceive and any concerns you have.” Vignette appeal and concern were measured on 5-point scales, as shown next.

Appeal

1. No Appeal
2. A Little Appeal
3. Some Appeal
4. Highly Appealing
5. Extremely Appealing

Concern

1. No Concern
2. A Little Concerning
3. Some Concern
4. Highly Concerning
5. Extremely Concerning

Participants were categorized as Promise when appeal ratings were greater than or equal to concern ratings and as Peril when concern ratings exceeded appeal ratings.

The analytic sample comprises the 229 respondents who answered the trust item (Q7); this criterion, rather than Qualtrics’s own completion flag, defines the analytic sample used throughout this paper. Qualitative response counts vary by item because missing responses were excluded from item-specific analyses. Responses were thematically coded using the same coding framework developed for the interview transcripts. Thirty-two final codes were applied across the dataset, capturing provider perspectives on trust, ethical concerns, bias, implementation feasibility, cost, health literacy, provider benefit, and patient engagement. These codes were subsequently grouped into thematic domains to support interpretation of patterns across the entire dataset. These broader thematic domains are revisited in the Discussion to contextualize the code-level findings presented in the Results below. Representative quotes from the survey are presented in the results section to illustrate key findings.

We stratified the frequency of the codes based on trust level (high vs. low) using two-proportion tests of significance. Because each table involves many simultaneous comparisons, we applied the Benjamini-Hochberg procedure to control the false discovery rate within each table (Table 3; Table 4; each vignette in Table 6 treated as its own family of comparisons). We report the raw p-value for each comparison in the tables and limit our discussion of trust-group differences in the narrative to codes that remained significant after this correction (Benjamini-Hochberg adjusted q ≤ 0.05).

## Results

### Study 1: Narrative Insights from Thought Leader Interviews

Analysis of the Study 1 interviews with healthcare organizational and policy leaders produced five recurring themes: trust in AI, ethical concerns and bias, financial and infrastructure constraints, digital literacy and patient engagement, and AI integration practices.

Trust emerged as a precondition for adoption rather than a byproduct of it. As one digital-care-platform executive put it, “Folks who are traditionally disadvantaged don’t trust a lot of structure because they’ve felt abused… You’ve got to align trust and comfort level with anybody who is using whatever we’re suggesting, not just point at people and say, I’ve got this done for you.” Leaders described trust as shaped by cultural, generational, and institutional factors as much as by technical reliability, and tied directly to whether oversight and accountability were visible to the communities being served.

Ethical concerns centered on the risk that AI could widen, rather than narrow, existing disparities. A rural healthcare executive warned that tools built for well-resourced systems (“Epic is built for large hospitals, and our smaller rural hospitals don’t have access to it”) could deepen a two-tier system, a concern echoed by a health policy expert who drew a direct parallel to the uneven early rollout of telehealth.

Financial and infrastructure barriers were described as foundational rather than incidental. Rural leaders emphasized that adoption depends on demonstrable near-term return on investment (“you have to be able to show the savings to get them to buy into it”), while basic connectivity gaps (“we still don’t have the technology capabilities for them to video into a counselor because the internet is so spotty”) were framed as a more immediate obstacle than any feature of the AI tool itself.

Digital literacy and patient engagement were treated as inseparable from trust. Leaders stressed that adoption succeeds only when tools build on patients’ existing comfort with everyday technology, most patients already carry a phone, and when community health workers are resourced to do the one-on-one work of onboarding skeptical patients.

Finally, leaders described AI integration as a governance and workflow problem as much as a technology one, calling for vendor-provider co-design (“having the vendors create a template and having the internal team discuss it… what is missing from this?”) rather than tools imposed without input from the systems that would use them. Together, these interviews frame responsible AI deployment in safety-net settings as contingent on trust-building, equity-aware design, adequate infrastructure investment, and collaborative governance, themes that directly shaped the vignette constructs and survey items used in Study 2.

### Study 2: Thematic Analysis of Provider Perceptions about AI in Healthcare

As described under Methodology above, Study 2 included semi-structured lines of questioning about how participants’ thought about:

- Their current level of trust in AI technologies to support healthcare delivery
- The ways they thought AI could specifically benefit rural and other safety-net populations in healthcare

Our analysis of these topics includes an overall view of the sample plus a stratification by current level of trust (low vs high).

#### Survey Respondent Characteristics

Table 1 summarizes the professional and organizational characteristics of survey respondents.

**Table 1.** Sample Characteristics of Survey Respondents. Presents the professional and organizational characteristics of survey respondents (N = 229): current role or title (Q3), organization type (Q1), and time in current position (Q4). Two respondents (0.9%) did not report a role or title (Not reported, under Current Role/Title).

| Characteristic | n | % |
| --- | --- | --- |
| **Current Role/Title** |  |  |
| Nurse (RN, LPN) | 58 | 25.3% |
| Physician (MD, DO) | 56 | 24.5% |
| Administrative Staff | 32 | 14.0% |
| Nurse Practitioner (NP) or Physician Assistant (PA) | 30 | 13.1% |
| Healthcare IT Professional | 6 | 2.6% |
| Allied Health Professional | 6 | 2.6% |
| Medical Technician | 6 | 2.6% |
| Researcher/Scientist in a Healthcare Setting | 3 | 1.3% |
| Graduate student healthcare practitioner | 2 | 0.9% |
| Medical Student | 1 | 0.4% |
| Not reported | 2 | 0.9% |
| Other clinical staff | 27 | 11.8% |
| **Total** | **229** | **100.0%** |
| **Organization Type** |  |  |
| Private company | 88 | 38.4% |
| Hospital or Medical Center | 65 | 28.4% |
| Non-profit | 24 | 10.5% |
| Government agency | 13 | 5.7% |
| University or Research Institution | 11 | 4.8% |
| Nursing Home or Long-Term Care Facility | 10 | 4.4% |
| Community Health Center | 7 | 3.1% |
| Other | 11 | 4.8% |
| **Total** | **229** | **100.0%** |
| **Time in Current Position** |  |  |
| Less than one year | 11 | 4.8% |
| 1-5 years | 75 | 32.8% |
| 6-10 years | 45 | 19.7% |
| More than 10 years | 98 | 42.8% |
| **Total** | **229** | **100.0%** |

Table 2 presents the overall distribution of respondent trust in AI technologies, the basis for the low trust / high trust stratification used throughout the analyses that follow.

**Table 2.** Distribution of Respondent Trust in AI Technologies. Presents the distribution of survey respondents (N = 229) by their reported level of trust in AI technologies to support healthcare delivery, based on the item “To what extent do you trust AI technologies to support healthcare delivery?” (1 = completely trust to 5 = completely distrust). The High trust and Low trust subtotals reflect the dichotomized grouping used for all subsequent stratified analyses in this paper (see Methods).

| Response to trust item | Scale value | n | % |
| --- | --- | --- | --- |
| Completely trust | 1 | 12 | 5.2% |
| Somewhat trust | 2 | 97 | 42.4% |
| **High trust (subtotal)** | **1–2** | **109** | **47.6%** |
| Neutral | 3 | 73 | 31.9% |
| Somewhat distrust | 4 | 38 | 16.6% |
| Completely distrust | 5 | 9 | 3.9% |
| **Low trust (subtotal)** | **3–5** | **120** | **52.4%** |
| Total | N/A | 229 | 100.0% |

#### Current Level of Trust

Table 3 presents the themes that emerged from a thematic analysis of open-ended survey data (25,36). It shows the frequency of the various themes influencing participants’ degree of trust in AI technologies for health care delivery among low trust and high trust groups.

**Table 3.** Issues Impacting Trust in AI in Health Care. Presents the themes that emerged from a thematic analysis of open-ended survey data (N = 211; low trust n = 109; high trust n = 102). The frequency of the various themes influencing participants’ degree of trust in AI technologies for health care delivery are shown for the total and among low trust and high trust groups. The table reports the raw p-value for each comparison; differences that remained significant after applying the Benjamini-Hochberg correction for the 34 comparisons in this table (adjusted q ≤ 0.05) are flagged in the Sig. column and discussed in the narrative. Responses could receive multiple codes, so column percentages sum to more than 100%.

| **Code** | **Total%** | **Low%** | **High%** | **p** | **Sig.** |
| --- | --- | --- | --- | --- | --- |
| Building Trust | 39.8% | 50.5% | 28.4% | 0.0011 | Yes |
| Data Bias and Accuracy | 25.6% | 33.0% | 17.6% | 0.0105 |  |
| Maintaining Data Security, Privacy & Anonymity | 13.7% | 12.8% | 14.7% | 0.6947 |  |
| Efficient and Effective Care | 13.3% | 3.7% | 23.5% | 0.0000 | Yes |
| Adequate Policy, Governance & Oversight | 9.0% | 8.3% | 9.8% | 0.6948 |  |
| Administrative Tasks | 8.1% | 2.8% | 13.7% | 0.0034 | Yes |
| Diagnostic Capabilities | 7.1% | 2.8% | 11.8% | 0.0109 |  |
| Need for Human in the Loop | 7.1% | 6.4% | 7.8% | 0.6881 |  |
| Accuracy of Data | 6.6% | 1.8% | 11.8% | 0.0038 | Yes |
| Automate Documentation | 6.6% | 2.8% | 10.8% | 0.0192 |  |
| Personalized Care | 5.2% | 7.3% | 2.9% | 0.1509 |  |
| Desire for Human Touch | 4.7% | 6.4% | 2.9% | 0.2344 |  |
| Decision Making | 4.3% | 4.6% | 3.9% | 0.8110 |  |
| Questioning if AI Is Needed | 4.3% | 7.3% | 1.0% | 0.0224 |  |
| Interplay Between AI and User | 3.8% | 1.8% | 5.9% | 0.1240 |  |
| Enhanced Health Info / Education Access | 2.8% | 2.8% | 2.9% | 0.9343 |  |
| Changing Provider Roles | 2.4% | 3.7% | 1.0% | 0.1993 |  |
| Revolutionize Infrastructure to Support | 2.4% | 0.9% | 3.9% | 0.1517 |  |
| Inadequate Representation in Training Data | 1.9% | 1.8% | 2.0% | 0.9466 |  |
| Cost and Hesitancy to Change | 1.4% | 1.8% | 1.0% | 0.6003 |  |
| Governance Structure | 1.4% | 1.8% | 1.0% | 0.6003 |  |
| Time Savings | 1.4% | 0.9% | 2.0% | 0.5224 |  |
| Enable Improved Treatment and Outcomes | 1.4% | 0.9% | 2.0% | 0.5224 |  |
| Increased Accessibility | 0.9% | 0.0% | 2.0% | 0.1419 |  |
| Workforce Displacement | 0.9% | 0.9% | 1.0% | 0.9624 |  |
| Care for the Marginalized | 0.5% | 0.0% | 1.0% | 0.3001 |  |
| Constant Need to Adapt to Trends | 0.5% | 0.0% | 1.0% | 0.3001 |  |
| Ethical AI | 0.5% | 0.0% | 1.0% | 0.3001 |  |
| Health Literacy and Trust | 0.5% | 0.0% | 1.0% | 0.3001 |  |
| Interplay: Language, Culture & Physicality | 0.5% | 0.0% | 1.0% | 0.3001 |  |
| Social Determinants of Health | 0.5% | 0.0% | 1.0% | 0.3001 |  |
| Workforce Availability, Training & Turnover | 0.5% | 0.0% | 1.0% | 0.3001 |  |
| Patient Triage | 0.5% | 0.0% | 1.0% | 0.3001 |  |
| **Other** | 16.1% | 11.9% | 20.6% | 0.0872 |  |

As shown in Table 3, the degree of trust in AI among low-trust respondents and high-trust respondents are influenced by different factors. Among low-trust respondents, over half (50.5%) cited building trust as a core issue, followed closely by data bias and accuracy (33.0%) and maintaining data security, privacy, and anonymity (12.8%). These participants expressed deep concern over the fairness, reliability, and safety of AI systems. In contrast, high-trust respondents were more likely to emphasize the practical benefits of AI: 28.4% mentioned building trust, while efficient and effective care was cited by 23.5%, significantly more than the 3.7% among low-trust respondents (p<0.001). High-trust respondents also more often cited diagnostic capabilities (11.8%) and automating documentation (10.8%) as reasons for their confidence in AI. Additionally, high-trust respondents were more likely to mention accuracy of data (11.8% vs. 1.8%, p=0.004); the difference in adequate policy, governance, and oversight was not significant (9.8% vs. 8.3%).

Both groups identified building trust as the most frequently mentioned factor behind their current level of trust. (Note the high trust group includes a majority of respondents stating they have a trust level of “somewhat”.)

After applying the Benjamini-Hochberg correction for the 34 comparisons in this table, four differences remained significant. Low-trust respondents were more likely to cite building trust (50.5% vs. 28.4%, p=0.001). High-trust respondents were more likely to cite efficient and effective care (23.5% vs. 3.7%, p<0.001), administrative tasks (13.7% vs. 2.8%, p=0.003), and accuracy of data (11.8% vs. 1.8%, p=0.004). Data bias and accuracy (33.0% vs. 17.6% low-trust), questioning whether AI is needed (7.3% vs. 1.0% low-trust), diagnostic capabilities (11.8% vs. 2.8% high-trust), and automating documentation (10.8% vs. 2.8% high-trust) showed the same direction of difference in the uncorrected test (each p < 0.05) but did not remain significant once corrected for multiple comparisons. Notably, the themes that might be expected to divide the groups most sharply, maintaining data security, privacy and anonymity, adequate policy, governance and oversight, and the need for a human in the loop, did not differ significantly and were raised at similar rates by both.

In the course of our analysis of the qualitative data about trust, four themes emerged, which we identify and discuss below.

##### Building Trust

A central theme that emerged across responses was the foundational role of trust when integrating AI into healthcare. Providers consistently expressed skepticism, largely due to the relative novelty of the technology, the absence of long-term outcome data, and the ongoing need for human oversight.

> A graduate student healthcare practitioner in the high-trust group remarked, “There are not enough studies with long-term outcomes to prove AI is safe and effective,” pointing to the limited empirical validation of AI tools in clinical practice.

Even among those open to its potential, there was strong consensus on maintaining human authority in medical decision-making.

> One nurse firmly stated, “AI in no way should diagnose or recommend any medical guidance,” emphasizing that clinical decisions should remain with licensed professionals.

In some clinical situations like radiology, for example, AI’s ability for diagnostics is the equivalent to or even better than human professionals. Still, human experts are required to confirm diagnosis, communicate the diagnosis to colleagues, and deploy emotional intelligence and context awareness in working with patients to develop a viable treatment plan (37).

##### Data Bias and Accuracy

Concerns about data bias and accuracy were also prominent, particularly among those who expressed conditional trust in AI.

> A physician explained, “I trust that AI technology can sift through data… but I do not trust the results 100% since there are many examples of either confabulation or incorrect clinical identification,” highlighting the tension between data processing capabilities and clinical reliability.

These concerns were even more pronounced among low-trust respondents.

> One physician specifically pointed out, “The data AI technology is using is not diverse and inclusive,” underscoring the risks of algorithmic bias and the need for datasets that reflect diverse patient populations to ensure equitable care.

As we develop deeper knowledge of health AI systems and how to deploy them effectively, the definition of diverse patient populations will require additional thought. For example, the deployment of AI in a rural setting will need to be highly sensitive to the context specific features and lived experiences that define life in a particular rural setting. This means that developing algorithmic solutions that are context aware and culturally sensitive will require approaches to governance that are not fully established.

##### Maintaining Data Security, Privacy, and Anonymity

Maintaining data security, privacy, and anonymity emerged as another significant determinant of trust, with concerns spanning both high- and low-trust groups.

> A nurse noted, “The recent media about AI compromising people’s privacy online makes me a bit wary,” capturing a broader apprehension around digital vulnerability.

Even among high-trust participants, the need for formal safeguards remained essential.

> As one physician emphasized, “I need to know that the AI is going to be HIPAA compliant. The platform it is on also impacts whether I feel safe with it.”

Together, these perspectives signal that trust is inseparable from strong data governance and transparency around privacy protections.

##### Efficient and Effective Care

High-trust respondents more often emphasized the efficiency and effectiveness of AI in improving healthcare delivery.

> A nurse described AI as “very effective and sufficient in aiding part of our medical system and staff,” illustrating its perceived ability to streamline operations.
>
> Another physician explained how AI could “integrate records and abstract [sic] important information,” noting its utility in managing documentation, generating differential diagnoses, and supporting treatment planning.

However, low-trust perspectives reveal concern that the pursuit of efficiency may come at a cost to patient care.

> As one physician cautioned, “Feel like there is going to be a push from higher ups (whether administration or Medicare/Medicaid) to use AI to cut costs and it will create problems in medicine,” highlighting that perceived administrative motives may undermine trust in AI’s implementation, particularly when efficiency is prioritized over clinical judgment.

#### Perceived Benefits of Health AI for Safety Net and Rural Populations

Participants identified a variety of ways in which AI could positively impact care delivery for underserved and rural communities. Table 4 summarizes the most frequently mentioned themes across trust groups, offering a comparative view of how low- and high-trust respondents envision AI contributing to safety-net care.

**Table 4.** Perceived Ways Health AI Could Benefit Rural and Safety-Net Populations. Presents the themes that emerged from a thematic analysis of open-ended survey responses to the item asking respondents to identify specific ways AI could benefit rural and other safety-net populations (N = 196; low trust n = 98; high trust n = 98). The frequency of the various themes is shown for the total sample and among low trust and high trust groups. The table reports the raw p-value for each comparison; none of the 25 comparisons in this table remained significant after applying the Benjamini-Hochberg correction (adjusted q ≤ 0.05). Responses could receive multiple codes, so column percentages sum to more than 100%.

| Code | Total % | Low % | High % | p | Sig. |
| --- | --- | --- | --- | --- | --- |
| Increased Accessibility | 45.9% | 45.9% | 45.9% | 1.0000 |  |
| Efficient and Effective Care | 23.5% | 22.4% | 24.5% | 0.7361 |  |
| Administrative Tasks | 19.4% | 17.3% | 21.4% | 0.4699 |  |
| Decision Making | 16.8% | 19.4% | 14.3% | 0.3399 |  |
| Care for the Marginalized | 11.7% | 12.2% | 11.2% | 0.8244 |  |
| Diagnostic Capabilities | 11.7% | 11.2% | 12.2% | 0.8244 |  |
| Personalized Care | 11.2% | 8.2% | 14.3% | 0.1746 |  |
| Enable Improved Treatment and Outcomes | 10.2% | 9.2% | 11.2% | 0.6370 |  |
| Enhanced Health Info / Education Access | 9.2% | 11.2% | 7.1% | 0.3225 |  |
| Lack of Time, Funds, and Resources | 6.6% | 7.1% | 6.1% | 0.7741 |  |
| Telehealth / Telemedicine | 6.1% | 1.0% | 11.2% | 0.0029 |  |
| Patient Triage | 5.6% | 5.1% | 6.1% | 0.7563 |  |
| Social Determinants of Health | 4.1% | 4.1% | 4.1% | 1.0000 |  |
| Automate Documentation | 3.6% | 2.0% | 5.1% | 0.2482 |  |
| Interplay Between AI and User | 3.6% | 5.1% | 2.0% | 0.2482 |  |
| Constant Need to Adapt to Trends | 2.0% | 3.1% | 1.0% | 0.3123 |  |
| Revolutionize Infrastructure to Support | 1.5% | 2.0% | 1.0% | 0.5607 |  |
| Data Bias and Accuracy | 1.0% | 1.0% | 1.0% | 1.0000 |  |
| Health Literacy and Trust | 1.0% | 1.0% | 1.0% | 1.0000 |  |
| Intense and Regulated Industry | 1.0% | 1.0% | 1.0% | 1.0000 |  |
| Changing Provider Roles | 0.5% | 1.0% | 0.0% | 0.3161 |  |
| Cost and Hesitancy to Change | 0.5% | 1.0% | 0.0% | 0.3161 |  |
| Maintaining Data Security, Privacy & Anonymity | 0.5% | 1.0% | 0.0% | 0.3161 |  |
| Workforce Displacement | 0.5% | 1.0% | 0.0% | 0.3161 |  |
| Other | 13.8% | 14.3% | 13.3% | 0.8358 |  |

Table 4 depicts how respondents envision AI directly benefiting rural and safety-net populations. Increased Accessibility was by far the most frequently cited benefit, mentioned in 45.9% of responses, and this view was shared identically across trust groups (45.9% low trust vs. 45.9% high trust), suggesting a near-universal recognition that AI’s core value for these populations lies in expanding access to care. Efficient and effective care (23.5%) and administrative task support (19.4%) followed as secondary themes. Telehealth/telemedicine showed the largest gap by trust group, cited more often by high trust respondents (11.2%) than low trust respondents (1.0%; p=0.003), echoing the same pattern observed in Table 3, though this difference did not remain significant after correcting for the 25 comparisons in this table.

Unlike the trust-antecedent themes in Table 3, perceived benefits for rural and safety-net populations were strikingly consistent across trust groups: of the twenty-five codes in Table 4, none differed significantly by trust group after applying the Benjamini-Hochberg correction. Telehealth/telemedicine showed the largest uncorrected gap (1.0% low trust vs. 11.2% high trust, p=0.003) but did not remain significant once corrected. The subsections below therefore describe the content of each theme rather than differences between groups.

##### Increased Accessibility (Top code, shared across trust groups)

Across both trust groups, respondents most often framed AI’s benefit for rural and safety-net populations in terms of expanding access to care that would otherwise be difficult to obtain.

> A low-trust nurse practitioner explained, “If AI can provide more access to patient, who otherwise would have no access. If AI were to provide appropriate and factual information to be used and evaluated by a practitioner who uses critical thinking skills to agree or disagree with plan and or diagnosis.”
>
> A high-trust nurse practitioner elaborated, “Rural populations have less access and physical, sometimes financial barriers to see specialists. Thus, primary care could benefit from having a safety net and catching missing screenings or other trends in patient health. Having a personalized plan may help those with lower health literacy to participate in their health care.”
>
> A low-trust administrative staff member linked this directly to equity: “These populations are completely underserved and pay a penalty in terms of outcomes. Good AI tools give us a chance to improve health equity.”
>
> A high-trust physician offered a similar assessment: “With the inadequate care rural populations receive mainly to the ability to access a provider, AI has to be helpful.”

Enable Improved Treatment and Outcomes (10.2%) was a related but less frequently cited benefit, reflecting a smaller but consistent emphasis on AI’s potential to improve clinical outcomes directly, distinct from the more dominant theme of expanding access itself.

##### Efficient and Effective Care (Second most cited, consistent across trust groups)

The second most frequently cited benefit, mentioned by roughly a quarter of respondents in both trust groups, was AI’s potential to deliver more efficient and effective care.

> A nurse echoed this sentiment, explaining, “Patients want care and they want it fast. They want answers and to feel better. AI would assist in all of these things,” highlighting how timeliness and responsiveness are central to patient needs and may be supported by AI-driven tools.
>
> By contrast, a low-trust physician noted, “patients would perceive it as a benefit only, if a noticeable care change affects them.,” underscoring the conditional nature of acceptance in settings where skepticism remains high.

##### Administrative Tasks

Administrative burden reduction was another key theme, particularly among high trust respondents. The use of AI-based automative systems to support administrative tasks has been one of the earliest and most frequent applications in healthcare (9,38,39). Tasks that are routine in nature have been most amenable to automation. Moreover, the significant time expenditure on administrative tasks like clinical notes, billing, and management of electronic health records contributes to high levels of stress and burnout among healthcare practitioners.

> A nurse practitioner stated that “AI is likely an increasing tool in many services to include healthcare. It may help to streamline care and reduce administrative tasks,” suggesting that automation could help free up provider time for direct patient care.
>
> One school-based nurse in the low trust group expressed caution, stating, “I’m not exactly sure how AI fits into nursing care in a public-school setting. It’s a busy place and I can see the appeal of AI to help with administrative tasks, but privacy concerns outweigh the potential benefit, in my opinion.”

This view illustrates that for some low-trust respondents, the perceived risks, particularly around privacy, may still overshadow the administrative advantages of AI.

##### Decision Making

Decision support was cited more frequently by low trust respondents, indicating that while they recognized AI’s potential to assist with clinical judgments, they remained cautious.

> A nurse practitioner emphasized the need for more evidence and security, stating, “More information [is] needed to improve confidence in AI. Plus, there remains a lack in tight electronic information security.”

These responses reveal that while decision-making support is valued, it must be accompanied by safeguards and transparency to be embraced in lower trust environments. By contrast, high trust respondents conveyed greater confidence in AI’s role in improving care through better decision-making.

> One provider stated, “AI has the potential to significantly improve the precision and effectiveness of healthcare. By analyzing vast amounts of data, AI can help in early detection of diseases, personalize treatment plans, and provide continuous monitoring and support.”

##### Personalized Care

Personalized care was more commonly emphasized by high trust respondents, who viewed AI as a way to enhance, not replace, individualized medicine.

> A nurse practitioner explained that “some patients, especially under 40, may better understand and appreciate the personalization,” pointing to generational differences in comfort with technology.
>
> Even among low trust respondents, there was some openness to this potential, with an administrative staff member noting, “It will create an easier process if things are artificially modified to what actually fits every patient’s needs.”

These responses reflect cautious optimism that AI, when properly tailored, can support more patient-centered approaches.

##### Telehealth/Telemedicine (Largest trust-group gap, not significant after correction)

Telehealth and telemedicine showed the largest uncorrected trust-group gap of any code in Table 4, more often emphasized by high trust respondents, though the difference did not remain significant once corrected for multiple comparisons; this may still reflect a modest trend toward greater enthusiasm for technology-enabled remote care.

> A clinical staff member noted, “We use AI in our everyday use now, and in most cases have had a better experience. So why would we not want that for our healthcare?”

Their statement suggests normalization of AI in other aspects of life has helped lay the groundwork for acceptance in medical contexts.

> However, caution remained among some low trust providers; one physician commented, “Patients are open to being seen sooner and getting the services they need in a timely manner. However, they need to be informed that using AI comes with risks.”

This remark highlights a recurring theme throughout the study: trust must be earned through transparency, validation, and meaningful patient education.

### Study 2: Analysis of Provider Reactions to Specific Health AI Tools

#### Quantitative Appraisal of Specific AI Tools

Figure 1 shows the mean appeal and concern rating for each vignette, including the 95% confidence interval (CI). Across all AI concepts/tools, the average appeal is greater than the average level of concern. To formally test these patterns while accounting for the repeated-measures design, in which each respondent rated all four vignettes, and for the trust segmentation used throughout this paper, Appeal and Concern ratings were each modeled with a random-intercept linear mixed model (Rating ∼ Vignette × Trust Group + (1 | Respondent); lme4::lmer, restricted maximum likelihood, Satterthwaite-approximated degrees of freedom).

**Figure 1.**
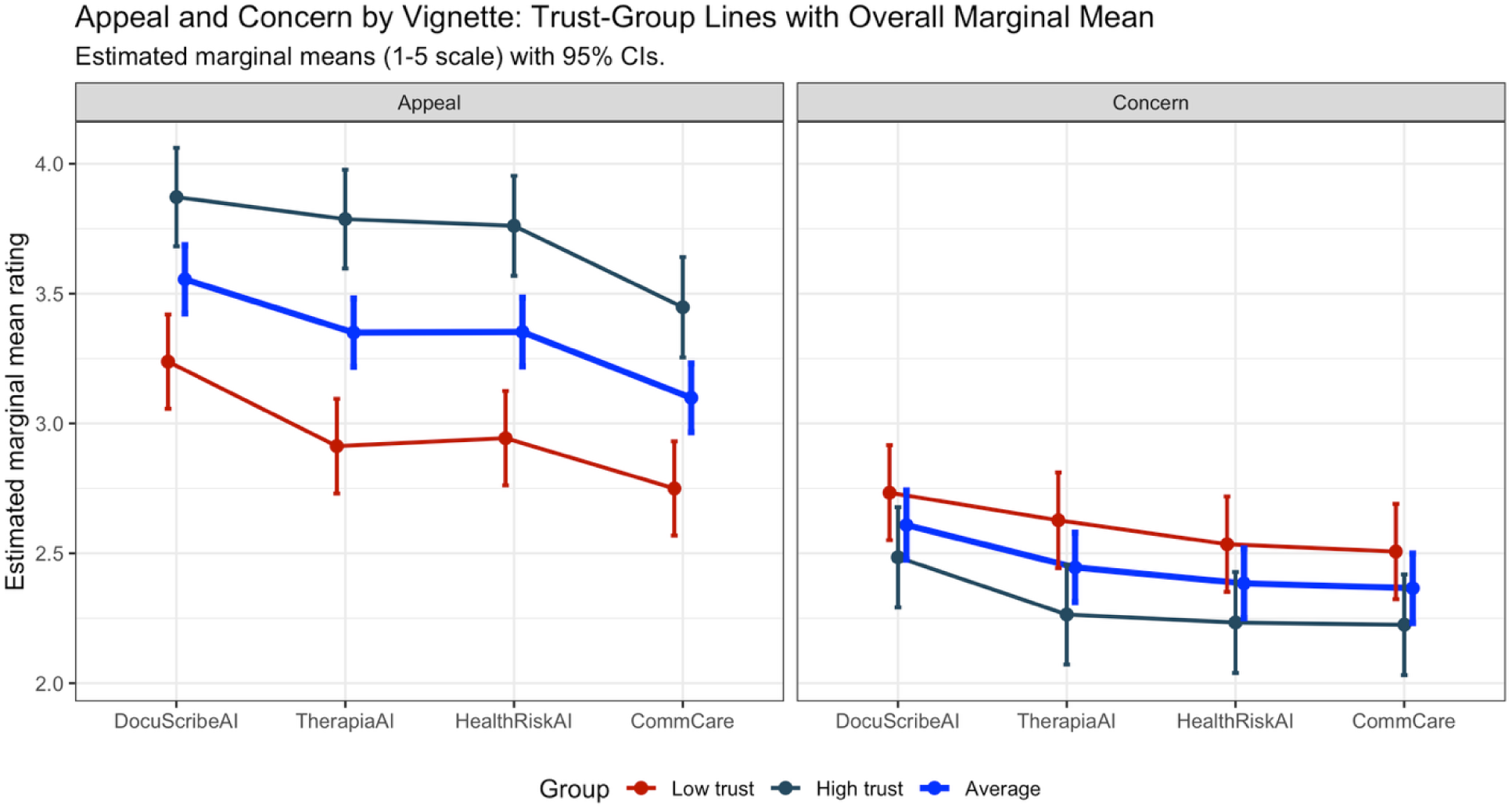
Average Appeal and Concern Rating for Each Vignette

Table 5 reports the estimated marginal mean rating for each vignette by trust group, along with the significance of the low-trust versus high-trust contrast within each vignette. Formal pairwise comparisons among the four vignette-estimated marginal means (Tukey-adjusted, from the same random-intercept mixed models) support this pattern statistically.

**Table 5.** Estimated Marginal Means for Vignette Appeal and Concern Ratings by Trust Group. Presents estimated marginal means, on the underlying 5-point scale (1 = No Appeal/No Concern to 5 = Extremely Appealing/Extremely Concerning), from random-intercept mixed models fit separately for Appeal and Concern, with vignette, trust group, and their interaction as fixed effects and a random intercept for respondent. Sig. reflects the significance of the low-trust versus high-trust contrast within each vignette. The vignette × trust group interaction was not significant for either outcome (all p > .08), indicating a consistent trust effect across vignettes.

| Vignette | Appeal:<br>Low trust | Appeal:<br>High trust | Appeal:<br>Sig. | Concern:<br>Low trust | Concern:<br>High trust | Concern:<br>Sig. |
| --- | --- | --- | --- | --- | --- | --- |
| DocuScribeAI | 3.24 | 3.87 | <0.001 | 2.73 | 2.48 | 0.067 |
| TherapiaAI | 2.91 | 3.79 | <0.001 | 2.63 | 2.26 | 0.008 |
| HealthRiskAI | 2.94 | 3.76 | <0.001 | 2.54 | 2.23 | 0.027 |
| CommCare | 2.75 | 3.45 | <0.001 | 2.51 | 2.22 | 0.039 |

For Appeal, DocuScribeAI was rated significantly higher than TherapiaAI (difference = 0.21, p = .016), HealthRiskAI (difference = 0.20, p = .018), and CommCare (difference = 0.46, p < .001). TherapiaAI and HealthRiskAI did not differ from each other (difference < 0.01, p > .99), but both were rated significantly higher than CommCare (differences of 0.25, p = .002 for each), which had the lowest appeal of the four tools.

For Concern, DocuScribeAI was again the outlier: rated significantly more concerning than TherapiaAI (difference = 0.16, p = .048), HealthRiskAI (difference = 0.22, p = .002), and CommCare (difference = 0.24, p < .001), while TherapiaAI, HealthRiskAI, and CommCare did not differ from one another on concern (all p > .58). DocuScribeAI was therefore simultaneously the most appealing and the most concerning of the four vignettes, while CommCare was both the least appealing and statistically indistinguishable from TherapiaAI and HealthRiskAI on concern.

#### Thematic Analysis of Provider Reactions to Specific Health AI Tools

Participants were also asked, for each vignette: “In a sentence or two, tell us a little more about the appeal you perceive and any concerns you have.” Responses were analyzed thematically using the same coding scheme applied elsewhere in this study. The same handful of themes recurred across all four tools, so Table 6 consolidates the qualitative results into a single cross-vignette table showing the total-sample percentage of respondents citing each code, rather than four separate tables; the full low-trust/high-trust breakdown for every code, none of which differed significantly by trust group after correction, is provided in the Supplement.

**Table 6.** Reasons for Appeal and Concern Across the Four AI Vignettes. Presents all nineteen codes cited in open-ended responses about each vignette’s appeal and concerns, as the total-sample percentage of respondents who cited each code. No code differed significantly by trust group in any of the four vignettes after applying the Benjamini-Hochberg correction (adjusted q ≤ 0.05); the full low-trust/high-trust breakdown, with per-code significance testing, is provided in S1 Text (Tables C-F).

| Code | DocuScribeAI | TherapiaAI | HealthRiskAI | CommCare |
| --- | --- | --- | --- | --- |
| Need for Human in the Loop | 29.7% | 18.5% | 11.9% | 2.6% |
| Accuracy of Data | 35.1% | 2.9% | 18.9% | 1.3% |
| Automate Documentation | 43.6% | 1.7% | 5.7% | 4.5% |
| Efficient and Effective Care | 9.9% | 16.2% | 11.3% | 21.8% |
| Time Savings | 32.2% | 8.1% | 5.0% | 6.4% |
| Maintaining Data Security, Privacy & Anonymity | 12.9% | 8.7% | 11.3% | 7.7% |
| Questioning if AI Is Needed | 10.9% | 1.7% | 8.8% | 13.5% |
| Personalized Care | 1.0% | 18.5% | 2.5% | 6.4% |
| Building Trust | 9.9% | 10.4% | 0.0% | 0.0% |
| Interplay Between AI and User | 5.4% | 1.7% | 10.1% | 3.8% |
| Data Bias and Accuracy | 2.0% | 16.2% | 0.0% | 0.0% |
| Decision Making | 2.5% | 3.5% | 9.4% | 1.3% |
| Patient Communications | 0.0% | 0.0% | 0.0% | 14.7% |
| Enhanced Health Info / Education Access | 0.0% | 13.3% | 0.0% | 0.0% |
| Patient Triage | 0.0% | 0.6% | 12.6% | 0.0% |
| Adequate Policy, Governance & Oversight | 7.9% | 0.6% | 0.0% | 0.0% |
| Desire for Human Touch | 1.0% | 5.8% | 0.0% | 0.0% |
| Unequal Access to Technology / Infrastructure | 1.0% | 0.0% | 0.6% | 3.2% |
| Other | 4.0% | 20.2% | 28.3% | 48.7% |

#### Cross-Vignette Themes

Two themes recur across every vignette regardless of trust group: the need to keep a human in the loop, and concern about data accuracy or bias.

##### Human in the loop

Need for human oversight is most prominent for the two tools with the most direct clinical stakes, DocuScribeAI and TherapiaAI, and least prominent for CommCare, but it does not differ by trust group for any of the four tools, suggesting a broad, trust-independent consensus that AI should support rather than replace clinical judgment.

As one nurse put it, discussing TherapiaAI, “there should always be a human reviewing or monitoring AI outputs; machines can’t replace clinical judgment and compassion. We can’t just rely on algorithms and hope for the best.”

##### Concern about data accuracy or bias

Concerns about accuracy carried a similar tone across tools;

One nurse, reflecting on DocuScribeAI, warned that “mistakes are made, words or medications [could be] heard incorrectly, which could cause major issues if not caught in a timely manner.”

Where trust group differences appear to emerge in the qualitative data, they echo the same direction already observed quantitatively in S1 Text (Tables C-F), though none reach significance once the Benjamini-Hochberg correction is applied within each vignette’s family of comparisons. Low-trust respondents showed a numerically higher rate of citing accuracy of data as a concern for HealthRiskAI (25.3% vs. 12.5% high-trust), of framing their views around building trust for TherapiaAI (16.1% vs. 4.7% high-trust), and of citing time savings as an appeal for CommCare (10.4% vs. 2.5% high-trust). High-trust respondents, in turn, showed a numerically higher rate of citing efficient and effective care for TherapiaAI (22.1% vs. 10.3% low-trust). None of these differences remained significant after correction. Data security, privacy, and administrative-burden themes were raised at similar rates by both trust groups across all four tools, indicating these are shared considerations rather than trust-dependent ones.

#### Vignette-Specific Themes

Two tool-specific codes stood out: Patient Triage for HealthRiskAI (12.6% of respondents) and Patient Communications for CommCare (14.7% of respondents), both directly address what those tools are designed to do. Their prominence, alongside a still-substantial “Other” category for HealthRiskAI and CommCare (28.3% and 48.7%, respectively), suggests provider reactions to these two newer, less clinically established tool concepts remain more heterogeneous and less settled than reactions to the more familiar DocuScribeAI and TherapiaAI concepts.

## Discussion

This study demonstrates that healthcare providers working in safety-net contexts are not passive recipients of AI technologies but active agents who negotiate the conditions under which AI is acceptable. Providers exhibit conditional optimism, supporting AI deployment when adequate governance, transparency, and human oversight are present, while expressing concern when these conditions are absent. Findings should be interpreted in light of the Texas-based sample and the organizational conditions reflected in these data.

This study explored healthcare providers’ perceptions of health AI in safety-net settings through three guiding research questions: (1) To what degree do providers trust health AI, and what shapes that trust? (2) How might health AI benefit safety-net populations, and does baseline trust moderate those perceived benefits? and (3) How do providers balance health AI’s benefits against their concerns, as explored through reactions to four specific hypothetical tools? Across these domains, a consistent pattern emerged: providers largely agree on what AI could do well and on what must be true before they trust it, and trust shapes the intensity of their optimism or caution more than it changes the substance of what they perceive. Both high- and low-trust respondents converged on the same top benefits for safety-net populations and the same non-negotiable conditions, preserving human oversight, ensuring data accuracy, and safeguarding privacy, for responsible adoption. Where trust did make a measurable difference was in degree: high-trust providers rated every hypothetical tool as more appealing and, for three of the four, less concerning, without changing which tool ranked where relative to the others.

### Trust in Health AI: Prevalence and Drivers (RQ1)

Roughly half of surveyed providers reported high trust in health AI (47.6%) and half reported low trust (52.4%), though this split is less polarized than it first appears: 60.8% of the low-trust group selected “neutral” rather than an actively distrusting response (Table 1).

Quantitatively, four factors distinguished high- and low-trust respondents after correcting for multiple comparisons (Table 2): low-trust respondents more often cited the need to build trust itself as a precondition (50.5% vs. 28.4%), while high-trust respondents more often cited efficient and effective care (23.5% vs. 3.7%), administrative task support (13.7% vs. 2.8%), and confidence in data accuracy (11.8% vs. 1.8%) as reasons for their current trust level. Notably, the factors that might be expected to divide the groups most sharply, data security and privacy, governance and oversight, and the need to keep a human in the loop, were raised at statistically indistinguishable rates by both groups. These appear to be baseline expectations nearly all providers hold regardless of their overall trust level, rather than points of disagreement between more and less trusting providers.

Study 1 interviews with health system and policy leaders add organizational-level texture to what shapes trust: demonstrable financial return on investment, baseline connectivity and technical infrastructure, and visible accountability to the communities served all emerged as preconditions leaders described for trust and adoption, alongside the individual-level factors identified in the survey.

### Perceived Benefits for Safety-Net Populations, and the Limited Role of Trust (RQ2)

When asked specifically how health AI could benefit rural and safety-net populations, providers across both trust groups converged overwhelmingly on a single answer: expanding access to care. Increased Accessibility was the most frequently cited benefit by a wide margin (45.9%), and it was cited at exactly the same rate by low-trust and high-trust respondents (45.9% each). Efficient and effective care (23.5%) and administrative task support (19.4%) followed as secondary, similarly shared benefits.

Of the twenty-five benefit-related codes analyzed, none differed significantly between trust groups after correcting for multiple comparisons, indicating that baseline trust in AI does not meaningfully moderate how providers perceive AI’s potential benefit to safety-net populations specifically. This is a notable contrast with RQ1’s finding that trust is differentiated by specific factors (efficient care, administrative support, data accuracy) when the question is about trust in AI generally: when the lens narrows to safety-net populations specifically, low- and high-trust providers reason about benefit in essentially the same way. Telehealth/telemedicine showed the largest raw gap (1.0% low-trust vs. 11.2% high-trust) but did not survive correction, a pattern worth revisiting with a larger sample given its plausible connection to broader technology enthusiasm.

This convergence suggests that skepticism about AI in general does not translate into a different vision of what AI could accomplish for underserved populations: providers who distrust AI today still see the same access-expanding potential as those who trust it; what differs, per RQ1, is their confidence that AI will deliver on that potential responsibly.

### Balancing Benefit and Concern: Evidence from Four Specific AI Tools (RQ3)

To examine how providers weigh benefit against concern for concrete, rather than abstract, AI applications, respondents rated their appeal and concern for four hypothetical tools: DocuScribeAI (clinical documentation), TherapiaAI (personalized treatment), HealthRiskAI (risk assessment and resource allocation), and CommCare (patient communication). Across all four, average appeal exceeded average concern (Figure 1), indicating that promise outweighs peril even at the level of specific applications, not just in the abstract.

Trust shaped the intensity but not the ranking of these reactions (Table 4). High-trust providers rated every tool as significantly more appealing than low-trust providers did (all p<.001), and rated three of the four tools (TherapiaAI, HealthRiskAI, CommCare) as significantly less concerning; DocuScribeAI was the exception, where concern did not differ significantly by trust group. Critically, the interaction between vignette and trust group was not significant for either appeal or concern, meaning trust raises or lowers the whole profile of reactions without changing which tool providers found most or least appealing, or most or least concerning. DocuScribeAI was simultaneously the most appealing and the most concerning of the four tools, a reminder that appeal and concern are not opposite ends of a single dimension but can rise together for a tool with high perceived clinical stakes and high perceived value. CommCare, by contrast, had the lowest appeal but was statistically indistinguishable from TherapiaAI and HealthRiskAI on concern, and produced the most heterogeneous open-ended reactions of the four tools (48.7% falling outside the study’s established coding scheme), suggesting provider views on this newer, less clinically established application remain the least settled (Table 5).

Qualitatively, providers balanced benefit against concern through two conditions that recurred across all four tools regardless of trust group: preserving a human in the loop, and ensuring the underlying data are accurate and unbiased. Neither concern differed significantly by trust group for any tool, reinforcing RQ1’s finding that these are shared, near-universal expectations rather than a fault line between more and less trusting providers. Providers’ vision for responsible deployment, then, is less about resolving disagreement between trust groups and more about institutionalizing the conditions, human oversight, data accuracy, and transparent governance, that both groups already agree are necessary, while remaining attentive to the greater interpretive uncertainty surrounding newer applications like HealthRiskAI and CommCare.

### Zooming Out: Cross-Cutting Thematic Domains

As introduced in Methods, the individual codes discussed throughout this study’s qualitative results also map onto five broader thematic domains: the performance, reliability, and clinical validity of AI tools; data governance, privacy, and security; the enhancement of clinical capability and care delivery; the human dimensions of AI in healthcare, including the desire for human touch and the interplay between AI and the people who use it; and the structural and organizational conditions that support or hinder adoption. Returning to our three research questions through this domain-level lens surfaces patterns that are not visible when each question is considered on its own terms.

For RQ1, trust drivers are dominated by the performance, reliability, and clinical validity domain (72.0% of coded mentions in Table 2), with a substantial secondary presence from the clinical capability and care delivery domain (31.8%), well ahead of the data governance, privacy, and security domain (22.3%). Providers’ trust judgments rest primarily on whether AI performs reliably, but nearly a third of the time also draw on whether it can meaningfully improve care, rather than on governance and privacy concerns specifically. RQ3 reinforces this: the two conditions that recurred across all four hypothetical tools regardless of trust group, preserving a human in the loop and ensuring data accuracy, are themselves both performance and reliability codes. Performance and reliability, in other words, function as the standing precondition providers apply whether the question is about trust in AI generally or reactions to a specific hypothetical tool.

RQ2 presents a more concentrated pattern still: perceived benefits for safety-net populations are almost entirely a clinical-capability-and-care-delivery phenomenon (90.8% of coded mentions in Table 4), a far higher concentration than trust drivers show for any single domain. This sharpens RQ2’s original finding that trust does not moderate benefit perception: it is not merely that high- and low-trust providers reach the same conclusion, it is that there is essentially one domain of reasoning in play regardless of trust level.

Placing these three domain-mappings side by side surfaces a pattern that no single research question, considered on its own, can show: the performance/reliability domain and the clinical-capability domain are the two domains that structure nearly all of this study’s qualitative data, appearing prominently, in some combination, across every question asked and every vignette tested. The performance/reliability domain ranges from 30.1% to 87.1% of coded mentions across the four vignette open-ends (S1 Text, Tables C-F), and the clinical-capability domain from 16.8% to 50.3%, while the remaining three domains, governance, human dimensions, and structural/organizational conditions, stay secondary throughout, never exceeding about a quarter of mentions for any single question or tool. Providers appear to reason about health AI along two largely question-independent axes: whether it is reliable and governable, and whether it can meaningfully expand what care can be delivered. Governance-specific, human-dimension, and structural or organizational concerns are consistently present in this dataset but never become the dominant lens through which these safety-net providers evaluate AI. We use this same domain structure to locate our findings within the broader literature below.

### Alignment and Tensions with Conventional Literature Narratives

We organize this comparison by the same five thematic domains used above, since they capture the axes along which our own qualitative data are structured and let us locate points of alignment and departure from prior literature within each.

However, while the literature often adopts a tone of caution or even skepticism, emphasizing barriers such as infrastructural inadequacy (3,13–17), digital illiteracy (8,23), and data governance challenges (5,9), our findings add nuance by capturing providers’ conditional optimism. Rather than expressing blanket resistance, many providers in our study articulated a willingness to adopt AI tools, especially if those tools are embedded within accountable governance frameworks, are user-centered in design, and include provider oversight. This pragmatism in response suggests a shift from the literature’s focus on why AI fails to reach vulnerable settings to how it might succeed, given the right conditions.

#### Theme A: AI Performance, Reliability & Clinical Validity

Provider perspectives in this study align with literature emphasizing the risk of biased or under representative training data in clinical AI (20), particularly relevant to safety-net populations who may be poorly represented in the datasets used to build clinical AI models. This concern was echoed by both low- and high-trust respondents in our study, and concentrated specifically in reactions to TherapiaAI, where 16.2% of respondents raised data-bias or representation concerns, compared to 2.0% or less for the other three vignettes (S1 Text, Tables C-F). Our trust-based segmentation also echoes emerging evidence that clinicians cluster into distinct subgroups with systematically different AI acceptance depending on experience and risk perception (40).

#### Theme B: Data Governance, Privacy & Security

While many studies have called for inclusive design and equitable AI development (2,6,32), our findings underscore the urgency of implementation mechanisms that reflect these values. Providers advocated for community-engaged governance models, better infrastructure support, and co-design with clinicians, moving beyond theoretical calls for fairness to practical strategies for trust-building and uptake. Governance and privacy concerns were present but secondary throughout our own data, raised in roughly a fifth of trust-driver mentions (22.3%, Table 2) and never the plurality concern for any single vignette (S1 Text, Tables C-F). Their vision challenges the literature to focus not only on design ethics but also on the day-to-day realities of deploying AI in fragile systems with limited staffing, high patient loads, and fragmented data ecosystems.

#### Theme C: Clinical Capability & Care Delivery Enhancement

While ethical concerns such as bias, transparency, and accountability dominate much of the academic discourse (5,9,28), our findings revealed that providers are simultaneously focused on implementation feasibility: issues like time savings, documentation reduction, and easing diagnostic burdens were emphasized, especially for tools like DocuScribeAI and HealthRiskAI, whose leading codes (Automate Documentation, 43.6%, and Time Savings, 32.2%, for DocuScribeAI; Accuracy of Data, 18.9%, and Patient Triage, 12.6%, for HealthRiskAI) closely track this domain. This indicates that providers are not just thinking about whether AI is ethical or fair; they are also weighing whether it helps them deliver better care under existing constraints.

#### Theme D: Human Dimensions of AI in Healthcare

Digital literacy and trust remain key concerns in both the literature and our findings. Prior studies stress that access to broadband is insufficient if users do not trust or understand digital tools (15,18); this literature also speaks to the structural-access concerns discussed below, we treat it here for its emphasis on user trust and comprehension specifically. Our provider interviews reinforce this view, with multiple respondents pointing to the labor-intensive process of building patient comfort with AI tools and the need for culturally and linguistically tailored onboarding processes. High-trust respondents went a step further, suggesting that AI could enhance equity and engagement if deployed with local buy-in and community engagement, which reflects a more empowered and proactive stance than the largely risk-averse posture seen in many systematic reviews (3,7,8).

#### Theme E: Structural & Organizational Conditions for Adoption

Prior work by Rajkomar et al. and Browning et al. highlights the integration and interoperability limitations in safety-net environments, particularly due to fragmented EHR systems and limited budgets (5,22). Our participants confirmed these constraints, yet many still expressed enthusiasm for tools like DocuScribeAI and HealthRiskAI, noting that such tools could reduce provider burnout and improve efficiency, if they are designed to integrate seamlessly into existing workflows and are supported by vendor-neutral platforms. These frontline insights extend the literature by emphasizing solution-focused strategies, such as tailoring AI for legacy EHR systems, using cloud-based deployments, and supporting training and change management within resource-limited clinics.

In sum, organizing this comparison by the same five thematic domains used to reflect on our research questions above shows that our findings both reaffirm long-standing literature concerns, particularly around data representativeness, infrastructural constraint, and inclusive design, and extend that literature by showing frontline safety-net providers as active agents negotiating these trade-offs rather than passive recipients of AI tools. Future research and policy efforts would benefit from centering these provider voices, organized around the same domains, to bridge the gap between theoretical calls for equitable AI and the day-to-day realities of deploying it in settings that, historically, have been overlooked and underserved.

## Limitations

This study has several limitations that warrant consideration. First, the sample, while diverse in terms of provider role, organization type, and tenure (Table 1), was limited to healthcare professionals in Texas, which may affect the generalizability of findings to safety-net settings in other geographic regions. Second, the trust coding used to segment respondents into “high-trust” and “low-trust” categories was rests on a single self-report item, groups neutral respondents with those expressing distrust, and may not capture the full nuance of individual perspectives. Third, although the mixed-methods design provided complementary insights, survey and interview data were not collected from the same participants, limiting the ability to directly connect responses. Additionally, social desirability bias may have influenced how some participants discussed their patients’ responses to AI or their institutional readiness. Finally, the study focused primarily on provider perspectives and did not incorporate the voices of patients themselves, which are essential for a more complete understanding of AI’s implications in safety-net care.

## Conclusion

As health AI tools continue to permeate clinical settings, their integration into safety-net healthcare systems presents both profound opportunities and critical risks. This study demonstrates that healthcare providers working in underserved environments are not categorically resistant to AI, rather, their perceptions are shaped by a nuanced evaluation of both promise and peril. While many express cautious optimism about AI’s potential to streamline workflows, personalize care, and reduce burnout, concerns remain acute regarding data integrity, ethical oversight, equity, and infrastructural readiness.

One of the study’s central findings is the importance of trust, not just in the technology itself, but in the institutions, data, and processes surrounding its development and deployment. High-trust providers tend to view AI as an augmentative force, capable of enhancing clinical decision-making and expanding care access, particularly through tools like DocuScribeAI and TherapiaAI. Low-trust providers, in contrast, are more attuned to systemic risks: the possibility of bias, the fragility of patient privacy, and the lack of resources to meaningfully support implementation. Despite these differences, there is strong alignment across both groups on the need for human-in-the-loop models and the imperative for AI tools to adapt to the realities of local practice environments. Providers emphasized that successful AI deployment depends not only on technical soundness but also on context-sensitive implementation rooted in cultural competence, clear governance, and community engagement.

This research contributes to the growing literature by highlighting how real-world practitioners are actively negotiating the trade-offs of health AI, balancing enthusiasm with critique and hope with caution. Rather than seeing safety-net providers as barriers to innovation, this study reveals them as critical architects of AI innovation and responsible deployment. Their insights offer a roadmap for developers, policymakers, and health systems alike. Moving forward, efforts to integrate AI into safety-net care must prioritize co-design with frontline providers, equitable infrastructure investment, and robust accountability mechanisms. Future research should expand to include patient perspectives, longitudinal evaluations of AI implementation, and analyses of policy levers that can enable ethical and inclusive adoption. Ensuring that Health AI fulfills its promise without reinforcing legacy disparities will require sustained collaboration across sectors and a commitment to centering the voices of those most affected by technological change.

## Data Availability

De-identified data may be made available from the corresponding author upon reasonable request.

## Declarations

Ethics and Consent: This study was reviewed by the Institutional Review Board of the authors’ institution and granted an exempt determination (STUDY00005660). All participants in this study gave consent for their participation, including permission to publish results from the study.

## Acknowledgments

Not applicable.

## Author Contributions

Conceptualization: CW, MKK. Funding acquisition: CW, MKK. Investigation: IP, MKK, ES, GP. Formal analysis: IP, MKK, ES, GP. Writing - original draft: IP, MKK. Writing - review and editing: IP, MKK, ES, GP, CW.

## Data Availability Statement

The data underlying the results presented in this study contain potentially identifying information about individual healthcare providers and are subject to the terms of the Institutional Review Board’s exempt determination (STUDY00005660). As such, they cannot be made publicly available. De-identified data may be made available from the corresponding author upon reasonable request.

## Clinical trial number

Not applicable.

## Supporting Information

**S1 Text. Reporting checklists, full vignette stimulus text, and vignette-level statistical results by trust group.** Contains the COREQ checklist for the Study 1 interviews (Table A), the CHERRIES checklist for the Study 2 survey (Table B), the full text of the four AI tool vignettes exactly as presented to survey respondents, and the complete low-trust/high-trust breakdown with significance testing for all nineteen open-ended-response codes for each of the four vignettes (Tables C-F). No code’s low-trust vs. high-trust difference reached significance in any vignette after Benjamini-Hochberg correction.

**S2 Dataset. Codebook: code definitions and illustrative quotations.** Lists each of the codes used in this study’s qualitative analysis, grouped by the five thematic domains discussed in the Discussion, with a plain-language description and up to three verbatim respondent quotations per code.

